# Quantifying SARS-CoV-2 transmission suggests epidemic control with digital contact tracing

**DOI:** 10.1101/2020.03.08.20032946

**Authors:** Luca Ferretti, Chris Wymant, Michelle Kendall, Lele Zhao, Anel Nurtay, Lucie Abeler-Dörner, Michael Parker, David Bonsall, Christophe Fraser

## Abstract

The newly emergent human virus SARS-CoV-2 is resulting in high fatality rates and incapacitated health systems. Preventing further transmission is a priority. We analysed key parameters of epidemic spread to estimate the contribution of different transmission routes and determine requirements for case isolation and contact-tracing needed to stop the epidemic. We conclude that viral spread is too fast to be contained by manual contact tracing, but could be controlled if this process was faster, more efficient and happened at scale. A contact-tracing App which builds a memory of proximity contacts and immediately notifies contacts of positive cases can achieve epidemic control if used by enough people. By targeting recommendations to only those at risk, epidemics could be contained without need for mass quarantines (‘lock-downs’) that are harmful to society. We discuss the ethical requirements for an intervention of this kind.

## Introduction

COVID-19 is a rapidly spreading infectious disease caused by the novel coronavirus SARS-COV-2, a betacoronavirus, which has now established a global pandemic. Around half of infected individuals become reported cases, and with intensive care support, the case fatality rate is approximately 2% (*1*). More concerning is that the proportion of cases requiring intensive care support is 5%, and patient management is complicated by requirements to use personal protective equipment and engage in complex decontamination procedures (*2*). Fatality rates are likely to be higher in populations older than in Hubei province (such as in Europe), and in low-income settings where critical care facilities are lacking (*3*). In ref. (*4*) the public health cost of failing to achieve sustained epidemic suppression was estimated as 250,000 lives lost in the next few months in Great Britain, and 1.1-1.2 million in the USA, even with the strongest possible mitigation action to ‘flatten the curve’. Even modest outbreaks will see fatality rates climb as hospital capacity is overwhelmed, and the indirect effects caused by compromised health care services have yet to be quantified.

No treatment is currently available, and a vaccine will not be available for several months (as of March 2020) at the earliest. The only approaches that we currently have available to stop the epidemic are those of classical epidemic control, such as case isolation, contact tracing and quarantine, physical distancing and hygiene measures.

The basic reproduction number R_0_ is the typical number of infections caused by an individual in the absence of widespread immunity. Once immunity becomes widespread, the effective reproduction number R will become lower than R_0_ and once R is less than 1, the population has herd immunity and the epidemic declines. Immunity can only safely be obtained by vaccination. Here we use the term “sustained epidemic suppression” to mean a reduction of the reproduction number R to less than 1 by changing non-immunological conditions of the population that affect transmission, such as social contact patterns.

The biological details of transmission of betacoronaviruses are known in general terms: these viruses can pass from one individual to another through exhaled droplets (*5*), aerosol (*6*), contamination of surfaces (*7*), and possibly through fecal-oral contamination (*8*). Here we compare different transmission routes that are more closely aligned to their implications for prevention. Specifically, we propose four categories:

I. *Symptomatic transmission:* direct transmission from a symptomatic individual, through a contact that can be readily recalled by the recipient.
II. *Pre-symptomatic transmission:* direct transmission from an individual that occurs before the source individual experiences noticeable symptoms. (Note that this definition may be context specific, for example based on whether it is the source or the recipient who is asked whether the symptoms were noticeable.)
III. *Asymptomatic transmission:* direct transmission from individuals who never experience noticeable symptoms. This can only be established by follow-up, as single time-point observation cannot fully distinguish asymptomatic from pre-symptomatic individuals.
IV. *Environmental transmission:* transmission via contamination, and specifically in a way that would not typically be attributable to contact with the source in a contact survey (i.e., this does not include transmission pairs who were in extended close contact, but for whom in reality the infectious dose passed via the environment instead of more directly). These could be identified in an analysis of spatial movements.

We acknowledge that boundaries between these categories may be blurred, but defined broadly these categories have different implications for prevention, responding differently to classical measures of case isolation and quarantining contacts (*9*) (*10*) (and for a specific application to COVID-19 see below (*11*)).

Evidence exists for each of these routes of transmission: symptomatic (*12*), pre-symptomatic (*13*); asymptomatic (*14*); and environmental (*12*). For prevention, the crucial information is the relative frequency of different routes of transmission so as to allocate finite resources between different intervention strategies.

Li *et al*. (*12*) presented self-reported data on exposure for the first 425 cases in Wuhan; some of these recorded visits to the Huanan Seafood Wholesale Market. The generalizability of transmission in that setting to other settings is highly uncertain, as this large-scale event seeded the epidemic in the absence of any knowledge about the disease. After closure of the Huanan Seafood Wholesale Market on January 1, 2020, of 240 cases with no exposure to any wet market, 200 individuals (83%) reported no exposure to an individual with respiratory symptoms. Inaccurate recall may explain some responses, including failing to notice symptoms were exceptional at a time before awareness of the disease began, but it is unlikely to be as much as 83% of them, implying that many individuals were infected by non-symptomatic individuals.

The situation in Singapore at first glance appears different, since unlike in Wuhan, many individuals were linked to an identified symptomatic source. However, the main difference is that the linkage was retrospective, such that linkage could be established even if transmission occurred before a case was symptomatic. As of March 5, 2020, there were 117 cases, of which 25 were imported. By devoting considerable resources, including police investigation, 75 of the 92 cases of local transmission were traced back to their presumed exposure, either to a known case or to a location linked to spread (*15*). Linking cases via a location generally includes the possibility of environmentally mediated transmission. Therefore, the large fraction of traceable transmission in Singapore does not contradict the large fraction without symptomatic exposure in Wuhan. However, it does suggest that transmission from asymptomatic, rather than pre-symptomatic, individuals is not a major driver of spread. Although serological surveys are currently lacking, other lines of evidence suggest that the scenario of many asymptomatic infections for each symptomatic one is unlikely. Testing 1,286 close contacts of confirmed cases found that, among 98 individuals testing positive, only 20% did not have symptoms at first clinical assessment (*16*). Among 634 individuals testing positive on board the Diamond Princess cruise ship, the proportion of individuals without symptoms was found to be 52%; the proportion who were asymptomatic (rather than pre-symptomatic) was estimated as 18% (*17*).

Testing of passengers on board six repatriation flights from Wuhan suggest that 40-50% of infections were not identified as cases (*18*) (*4*). Mild cases have been found to have viral loads 60-fold less than severe cases (*19*) and it is likely that the viral loads of asymptomatic individuals are lower still, with possible implications for infectiousness and diagnosis.

The most accurate and robust quantification of the relative frequency of routes of transmission would be a well-designed prospective cohort study with detailed journal and phylogenetic investigations.

However, the current global emergency requires timely estimates using imperfect data sources. We performed a detailed analysis of the timing of events in defined transmission pairs, derived the generation time distribution, and attributed a probability for each pair that transmission was pre-symptomatic. We also fit a mathematical model of infectiousness through the four routes discussed above, over the course of infection. This allowed us to calculate R_0_, estimate the proportion of transmission from different routes, and make predictions about whether contact tracing and isolation of known cases is enough to prevent spread of the epidemic.

### Estimating SARS-CoV-2 transmission parameters

We used the exponential growth rate of the epidemic, *r*, from the early stages of the epidemic in China, such that the effect of control measures discussed later will be relative to the early stages of an outbreak, exemplified by baseline contact patterns and environmental conditions in Hubei during that period. We note that this assumption is implicit in many estimates of R_0_. The epidemic doubling time T_2_ is equal to log_e_(2) / *r*. We used the value r = 0.14 per day (*20*), corresponding to a doubling time of 5.0 days.

The incubation period is defined as the time between infection and onset of symptoms. It is estimated as the time between exposure and report of noticeable symptoms. We used the incubation period distribution calculated by (*21*). The distribution is lognormal with mean 5.5 days, median 5.2 days and standard deviation 2.1 days, and is included with our results in Fig. 1.

**Fig. 1.**
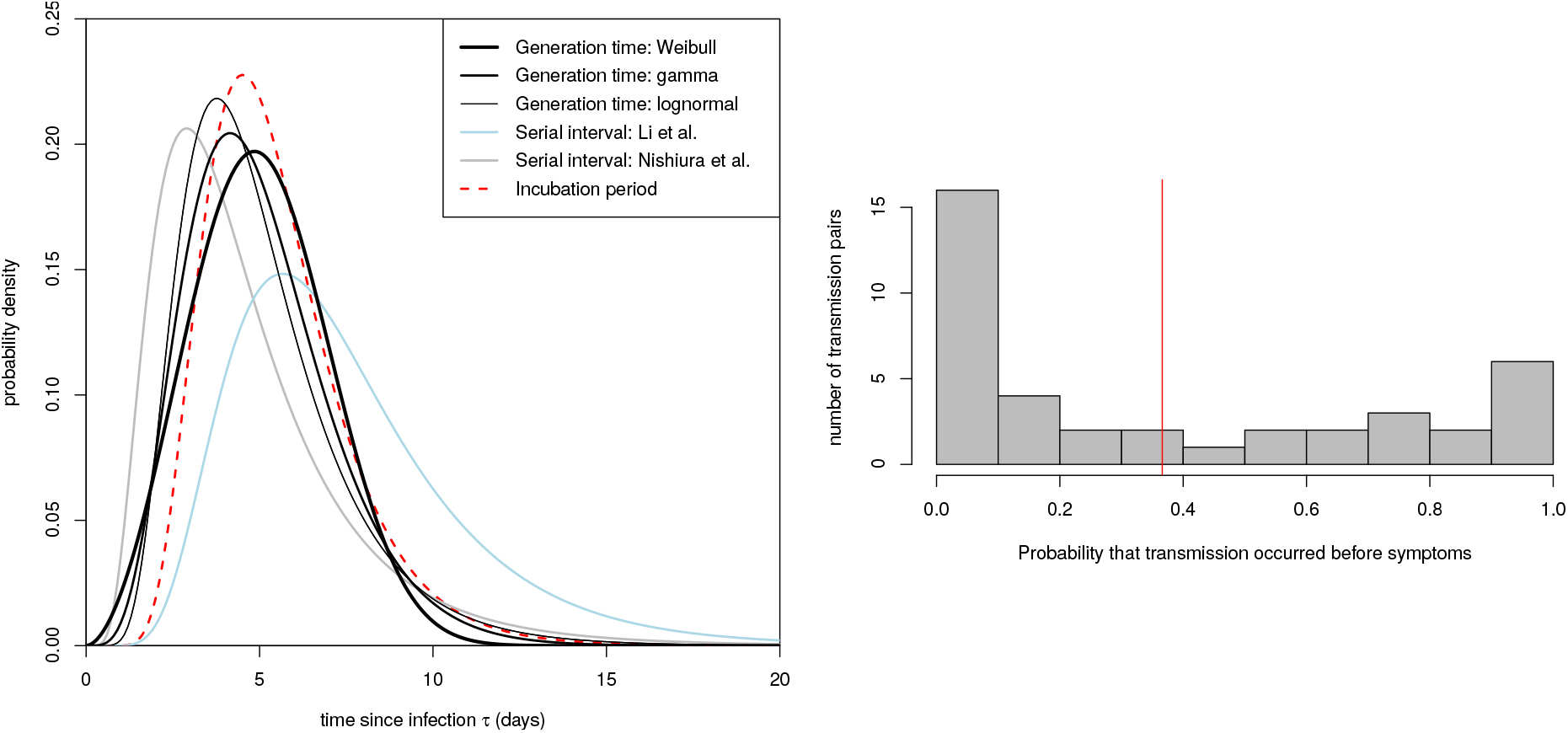
Quantifying transmission timing in 40 transmission pairs. Left panel: our inferred generation time distributions, in black; thicker lines denote higher support for the corresponding functional form, with the Weibull distribution being the best fit. For comparison we also include the serial interval distributions previously reported by (*12*) (in light blue) and (*22*) (in grey), and the incubation period distribution we used here from (*21*) (dashed red line). Right panel: the distribution of the posterior probability of pre-symptomatic transmission for each of the 40 transmission pairs. The red vertical line shows the mean probability.

The generation time is defined for source-recipient transmission pairs as the time between the infection of the source and the infection of the recipient. Because time of infection is generally not known, the generation time is often approximated by the serial interval, which is defined as the time between the onset of symptoms of the source and the onset of symptoms of the recipient. We did not take that approach here; instead, we directly estimated the generation time distribution from 40 source-recipient pairs. These pairs were manually selected according to high confidence of direct transmission inferred from publicly available sources at the time of writing (March, 2020), and with known time of onset of symptoms for both source and recipient. We combined dates of symptom onset with intervals of exposure for both source and recipient (when available) and the above distribution of incubation times and from these inferred the distribution of generation times. The distribution is best described by a Weibull distribution (AIC=148.4, versus 149.9 for gamma and 152.3 for lognormal distribution) with mean and median equal to 5.0 days and standard deviation of 1.9 days, shown in the left panel of Fig. 1. We also show the results of sensitivity analysis to different functional forms, and compare to two previously published serial interval distributions in refs (*22*) and (*12*). Uncertainty in the fit of the distribution is shown in Fig. *S*1. Our distribution is robust with respect to the choice of transmission events (Fig. *S*2). Correlation in the uncertainty between the inferred mean and standard deviation is shown in Fig. *S*3. The distribution of serial intervals for these pairs is shown in Fig. *S*4. The countries from which the transmission pair data was obtained are shown in Fig. *S*5.

For each of the 40 transmission pairs we estimated the posterior probability that transmission was pre-symptomatic, i.e., occurred before the onset of symptoms in the infector. We used a Bayesian approach with an uninformative prior (transmission before or after symptoms equally likely). The 40 probabilities inferred are shown in the right panel of Fig. 1; the mean probability is 37% (95% CI: 27.5% - 45%), which can be interpreted as the fraction of pre-symptomatic transmission events out of pre-symptomatic plus symptomatic transmission events. This mean probability over all pairs approximates our prior, but the bimodal distribution of individual probabilities in Fig. 1 shows that these are typically far from the prior, i.e., the data are strongly informative. Uncertainty in the value of this fraction is shown in Fig. *S*6. The value does not depend significantly on the choice of prior (Figs *S*7 and *S*8), functional form of the distribution of generation times (Figs *S*9 and *S*10), or on the choice of transmission events (Fig. *S*11).

### A general mathematical model of SARS-CoV-2 infectiousness

We use a mathematical formalism (*23*) that describes how infectiousness varies as a function of time since infection, τ, for a representative cohort of infected individuals. This includes heterogeneity between individuals, and averages over those individuals who infect few others and those who infect many. This average defines the function *β*(*τ*). Infectiousness may change with *τ* due to both changing disease biology (notably viral shedding) and changing contact with others. The area under the *β* curve is the reproduction number R_0_.

We decompose *β*(*τ*) into four contributions that reflect our categorization above, namely asymptomatic transmission, pre-symptomatic transmission, symptomatic transmission, and environmental transmission. The area under the curve of one of these contributions gives the mean total number of transmissions over one full infection, via that route - asymptomatic, pre-symptomatic, symptomatic or environmental - which we define to be R_A_, R_P_, R_S_ and R_E_ respectively. The sum of these is R_0_.

The mathematical form for *β*(*τ*) is:

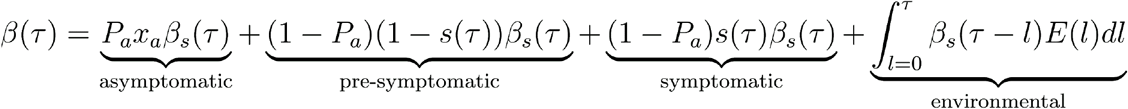

*β*_*s*_(*τ*) is the infectiousness of an individual currently either symptomatic or pre-symptomatic, at age-of-infection *τ*. Other parameters in this expression, and those feeding into it indirectly, are listed in Table 1. A detailed discussion of this expression including its assumptions is found in the supplementary materials. The priors chosen for parameters not directly calculated from data are shown in Supplementary Fig. *S*12. The infectiousness model result using central values of all parameters is shown in Fig. 2.

**Table 1:** Parameters of the infectiousness model

| Name | Symbol | Description | Central value | Uncertainty | Source |
| --- | --- | --- | --- | --- | --- |
| *Parameters directly calculated from data* |  |  |  |  |  |
| Doubling time | $T_2$ | The time taken for the epidemic to double in size during the early uncontrolled phase of expansion | 5.0 days | 95% CI 4.2 - 6.4 | (20) |
| Incubation period<br>(2 parameters) | $s(\tau)$ | lognormal meanlog<br>lognormal sdlog | 1.644<br>0.363 | 95% CI 1.495 - 1.798<br>95% CI 0.201 - 0.521 | (21) |
| Generation time<br>(2 parameters) | $w(\tau)$ | Weibull shape<br>Weibull scale | 2.826<br>5.665 | 95% CI 1.75 - 4.7<br>95% CI 4.7 - 6.9 | This paper |
| *Parameters with Bayesian priors informed by anecdotal reports or indirect evidence* |  |  |  |  |  |
| Proportion asymptomatic | $P_a$ | The proportion of infected individuals who are asymptomatic | 0.4 | Prior = beta($\alpha=1.5$, $\beta=1.75$)<br>Mode = 0.4<br>Mean = 0.46 | Media reports (Diamond Princess) |
| Relative infectiousness of asymptomatics | $x_a$ | The ratio of infectiousness of asymptomatic individuals to infectiousness of symptomatic individuals | 0.1 | Prior = beta($\alpha=1.5$, $\beta=5.5$)<br>Mode = 0.1<br>Mean = 0.21 | Observation of few missing links in Singapore outbreak to date. Suggestion from (19) |
| Fraction of all transmission that is environmentally mediated | $R_E/R_0$ | Self-explanatory | 0.1 | Prior = $\text{beta}(\alpha=1.5, \beta=5.5)$<br>Mode = 0.1<br>Mean = 0.21 | Anecdotal observation that many infections can be traced to close contacts once detailed tracing is completed. |
| Environmental infectiousness | $E(l)$ | Rate at which a contaminated environment infects new people after a time lag $l$ | 3 | Box function (0,n) days, Prior for $n = \text{Gamma}(\text{shape} = 4, \text{rate} = 1)$<br>Mode = 3<br>Mean = 4 | (24) - variety of values for many different surfaces. |

**Fig. 2.**
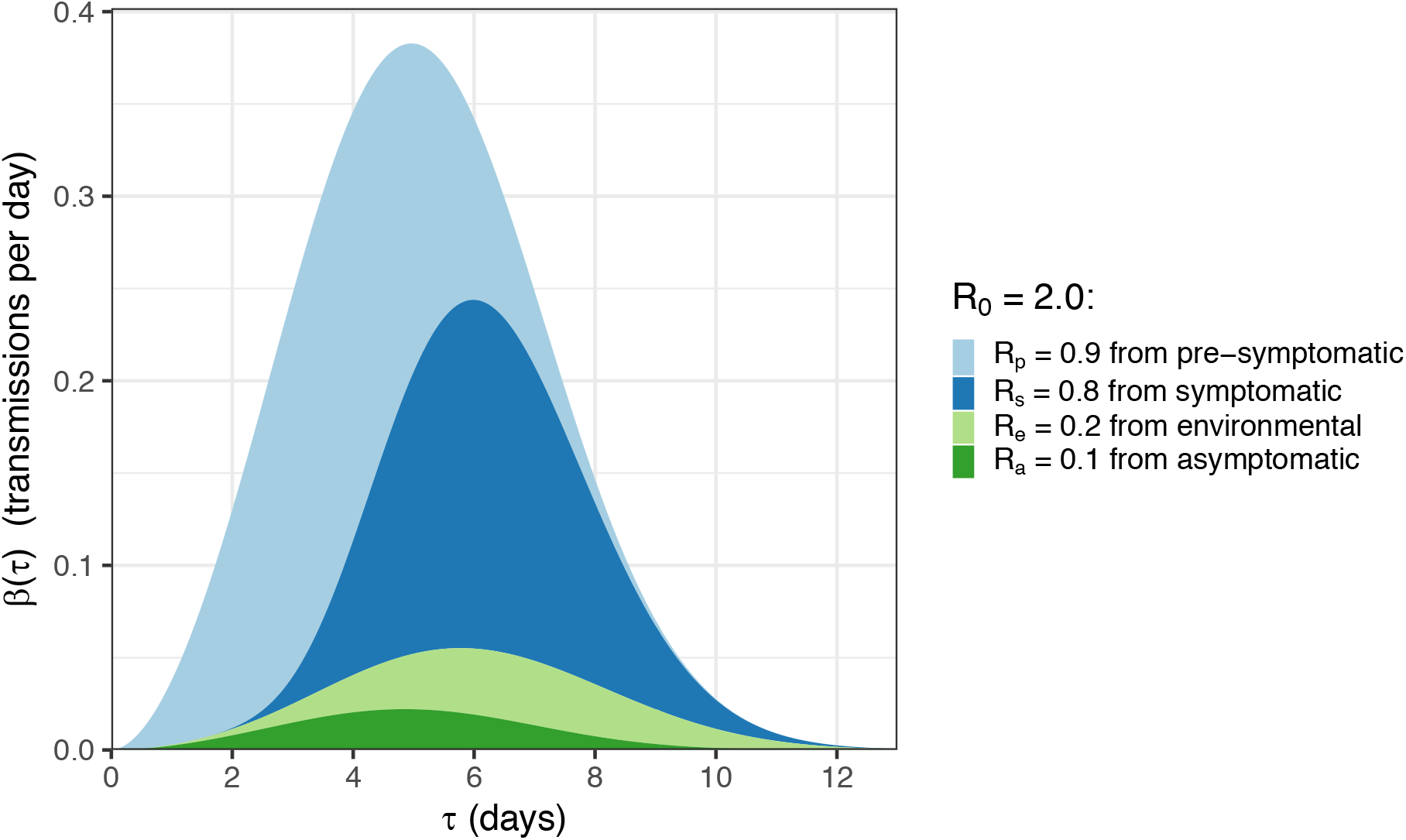
Our model of infectiousness. The average infectiousness (rate of infecting others), *β*, as a function of the amount of time since infection, *τ*. The total coloured area found between two values of *τ* is the number of transmissions expected in that time window. The total coloured area over all values of *τ* is the number of transmissions expected over the full course of one infection, i.e. the basic reproduction number R_0_. The different colours indicate the contributions of the four routes of transmission (stacked on top of one another), so that the total area of one colour over all values of *τ* is the average number of transmissions via that route over the whole course of infection: R_P_, R_S_, R_E_, and R_A_ for pre-symptomatic, symptomatic, environmentally mediated, and asymptomatic transmission respectively. Values are rounded to one decimal place. Stopping disease spread requires reduction of R to less than 1: blocking transmission, from whatever combination of colours and values of *τ* we can achieve, such that the total area is halved.

By drawing input parameter sets from the uncertainties shown in Table 1, we quantified our uncertainty in R_0_ and its four contributions. The resulting values are shown in Table 2 and their underlying distributions are shown in Fig. *S*13. Two-dimensional distributions showing correlations in uncertainty are shown in Fig. *S*14.

**Table 2:** R_0_ and its components.

| | Pre-symptomatic | Symptomatic | Environmental | Asymptomatic | Total $R_0$ |
| --- | --- | --- | --- | --- | --- |
| Absolute | Point estimate: 0.9<br>Uncertainty median: 0.7<br>CI: 0.2 - 1.1 | Point estimate: 0.8<br>Uncertainty median: 0.6<br>CI: 0.2 - 1.1 | Point estimate: 0.2<br>Uncertainty median: 0.4<br>CI: 0.0 - 1.3 | Point estimate: 0.1<br>Uncertainty median: 0.2<br>CI: 0.0 - 1.2 | Point estimate: 2.0<br>Uncertainty median: 2.1<br>CI: 1.7 - 2.5 |
| Fraction of $R_0$ | Point estimate: 0.47<br>Uncertainty median: 0.35<br>CI: 0.11 - 0.58 | Point estimate: 0.38<br>Uncertainty median: 0.28<br>CI: 0.09 - 0.49 | Point estimate: 0.1 by assumption<br>Uncertainty median: 0.19<br>CI: 0.02 - 0.56 | Point estimate: 0.06<br>Uncertainty median: 0.09<br>CI: 0.00 - 0.57 | 1 by definition |

The estimate of R_P_/(R_P_+R_S_) obtained by this method is 0.55 (0.37 - 0.72), which is larger than the estimate of 0.37 (0.28 - 0.45) from our analysis of the 40 transmission pairs but with overlapping uncertainty.

We define *θ* as the fraction of all transmissions that do not come from direct contact with a symptomatic individual: *1* − *RS* /*R0*. This corresponds to the *θ* of (*9*) in the case where there is only pre-symptomatic and symptomatic transmission. From Table 2 this is 0.62 (0.50 - 0.92). The value of *θ* observed during an exponentially growing epidemic will be distorted when the timing of the different contributions to transmission occur at different stages of the infection, due to over-representation of recently infected individuals. This effect can be calculated through use of the renewal equation, as was recently done to calculate the distribution of time from onset of COVID-19 symptoms to recovery or death (*20*) (see supplementary material). We calculated the *θ* that would be observed with the early exponential growth seen in China as 0.68 (0.56 - 0.92). The correction due to the epidemic dynamics is small compared to parameter uncertainties.

We developed our mathematical model of infectiousness into a web application where users can test the effect of alternative parameter combinations (*25*).

### Modelling case isolation and contract tracing with quarantine

We modelled the combined impact of two interventions: (i) isolation of symptomatic individuals, and (ii) tracing the contacts of symptomatic cases and quarantining them. These interventions aim to stop the spread of the virus by reducing the number of transmissions from both symptomatic individuals and from their contacts (who may not be symptomatic), while minimizing the impact on the larger population. In practice, neither intervention will be successful or possible for 100% of individuals. The success rate of these interventions determines the long-term evolution of the epidemic. If the success rates are high enough, the combination of isolation and contact tracing/quarantining could bring R below 1 and therefore effectively control the epidemic.

An analytical mathematical framework for the combined impact of these two interventions on an epidemic was previously derived in (*9*). In the supplementary material, we solve these equations using our infectiousness model above, i.e., quantifying how the SARS-CoV-2 epidemic is expected to be controlled or not by case isolation and the quarantining of traced contacts. Our results are shown in Fig. 3. The black line shows the threshold for epidemic control: combined success rates in the region to the upper right of the black line are sufficient to reduce R to less than one. The *x* axis is the success rate of case isolation, which can be thought of either as the fraction of symptomatic individuals isolated, assuming perfect prevention of transmission on isolation, or the degree to which infectiousness of symptomatic individuals is reduced assuming all of them are isolated. The *y* axis is the success rate of contact tracing; similarly, this can be thought of as the fraction of all contacts traced, assuming perfectly successful quarantine upon tracing, or the degree to which infectiousness of contacts is reduced assuming all of them are traced. These results for intervention effectiveness, and their dependence on all parameters in our combined analysis, can be explored through the same web interface as for our model of infectiousness (*25*).

**Fig. 3.**
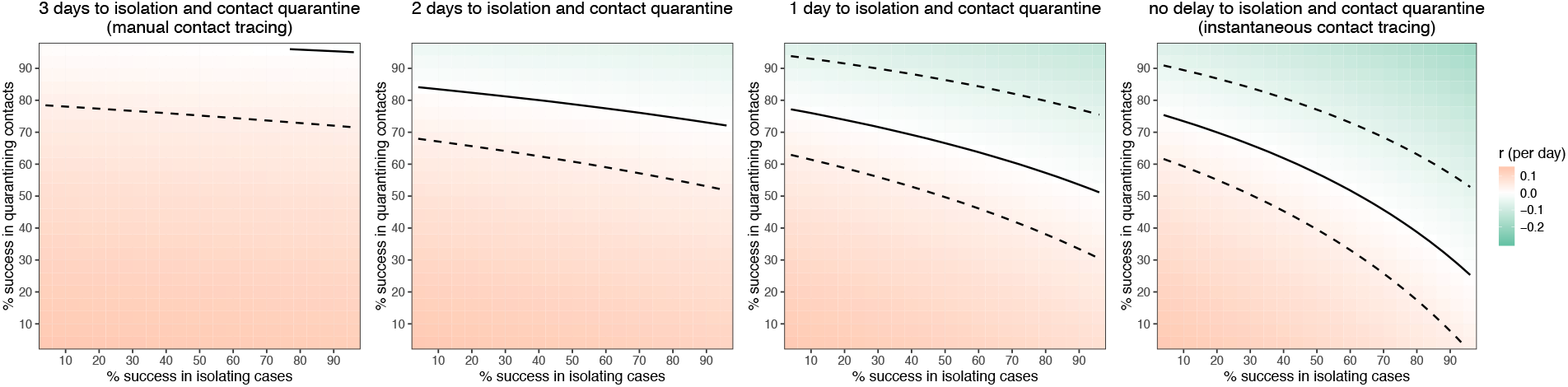
Quantifying intervention success. Heat map plot showing the exponential growth rate of the epidemic *r* as a function of the success rate of instant isolation of symptomatic cases (x axis) and the success rate of instant contact tracing (y axis). Positive values of *r* (red) imply a growing epidemic; negative values of *r* (green) imply a declining epidemic, with greater negative values implying faster decline. The solid black line shows *r*=0, i.e. the threshold for epidemic control. The dashed lines show uncertainty in the threshold due to uncertainty in R_0_ (see Supplementary Figures 15-17). The different panels show variation in the delay associated with the intervention - from initiating symptoms to case isolation and quarantine of contacts.

Delays in these interventions make them ineffective at controlling the epidemic (Fig. 3): traditional manual contact tracing procedures are not fast enough for SARS-CoV-2. A delay from confirming a case to finding their contacts is not, however, inevitable. Specifically, this delay can be avoided by using a mobile phone App.

### Epidemic control with instant digital contact tracing

A mobile phone App can make contact tracing and notification instantaneous upon case confirmation. By keeping a temporary record of proximity events between individuals, it can immediately alert recent close contacts of diagnosed cases and prompt them to self-isolate.

Apps with similar aims have been deployed in China. Public health policy was implemented using an App which was not compulsory but was required to move between quarters and into public spaces and public transport. The App allows a central database to collect data on user movement and coronavirus diagnosis and displays a green, amber or red code to relax or enforce restrictions on movement. The database is reported to be analyzed by an artificial intelligence algorithm that issues the colour codes (*26*). The App is a plug-in for the WeChat and Alipay Apps and has been generally adopted.

Mainland China outside of Hubei province received significantly more introductions from Wuhan than did anywhere else, following mass movements of people around Chinese New Year and the start of the Wuhan lockdown (*27*). Despite this, sustained epidemic suppression has been achieved in China: fewer than 150 new cases have been reported each day from March 2 to March 23, down from thousands each day at the height of the epidemic. South Korea has also achieved sustained epidemic suppression: 76 new cases on March 24, down from a peak of 909 on February 29, and is also using a mobile phone App for recommending quarantine. Both the Chinese and South Korean Apps have come under public scrutiny over issues of data protection and privacy.

With our result in Fig. 3 implying the need for extremely rapid contact tracing, we set out to design a simple and widely acceptable algorithm from epidemiological first principles, using common smartphone functionality. The method is shown in Fig. 4. The core functionality is to replace a week’s work of manual contact tracing with instantaneous signals transmitted to and from a central server.

**Fig. 4.**
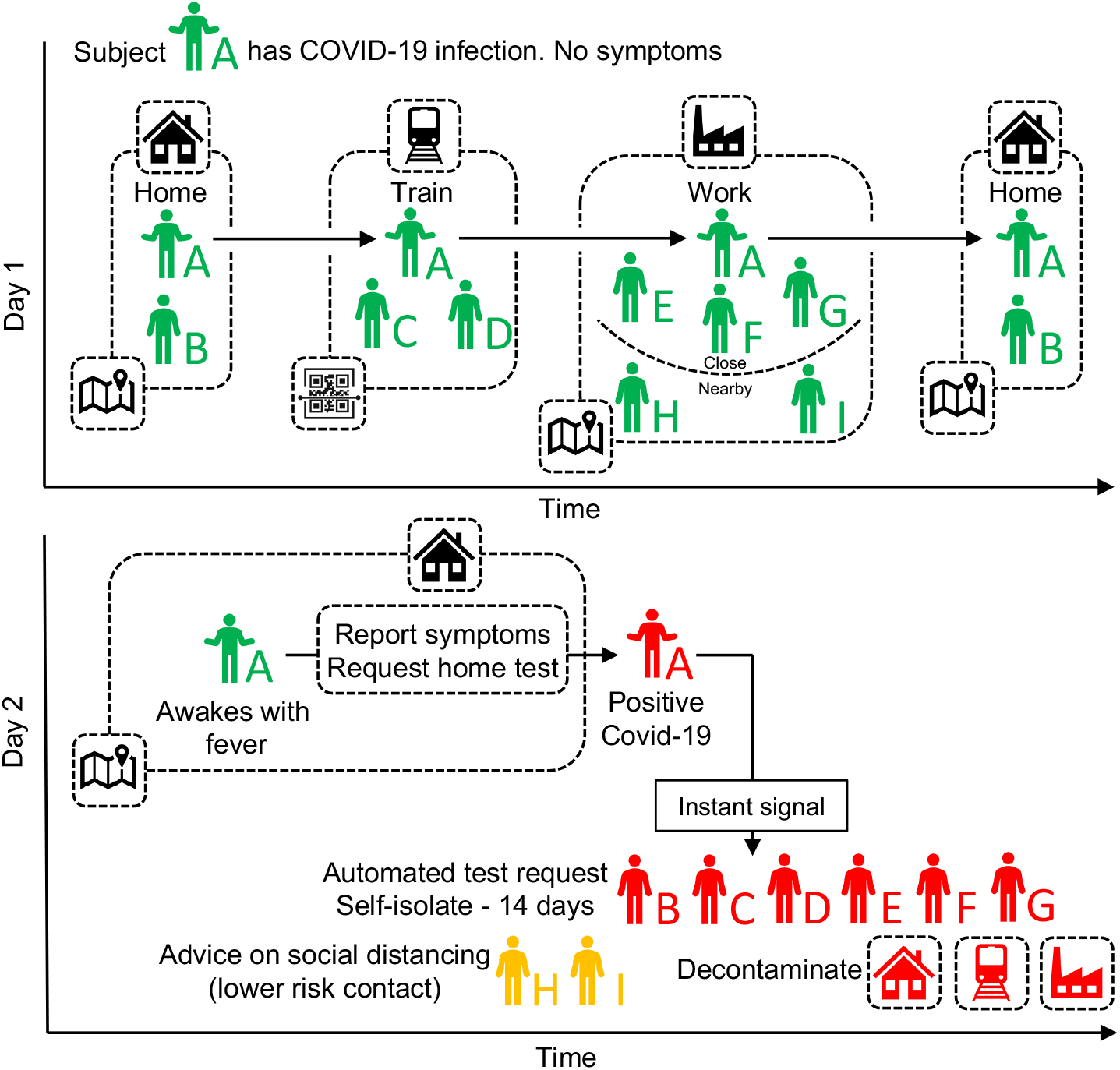
A schematic of app-based COVID-19 contact tracing. Contacts of individual A (and all individuals using the app) are traced using GPS co-localisations with other App users, supplemented by scanning QR-codes displayed on high-traffic public amenities where GPS is too coarse. Individual A requests a SARS-COV-2 test (using the app) and their positive test result triggers an instant notification to individuals who have been in close contact. The App advises isolation for the case (individual A) and quarantine of their contacts.

Coronavirus diagnoses are communicated to the server, enabling recommendation of risk-stratified quarantine and physical distancing measures in those now known to be possible contacts, while preserving the anonymity of the infected individual. Tests can be requested by symptomatic individuals through the App.

The simple algorithm can easily be refined to be more informative, for example quarantining areas if local epidemics become uncontrolled, quarantining whole households, or performing second- or third-degree contact tracing if case numbers are rising, which would result in more people being preemptively quarantined. Algorithmic recommendations can also be manually overridden, where public health officials gather more specific evidence. The App can serve as the central hub of access to all COVID-19 health services, information and instructions, and as a mechanism to request food or medicine deliveries during self-isolation.

In the context of a mobile phone App, Fig. 3 paints an optimistic picture. There is no delay between case confirmation and notification of contacts, leaving the total delay for the contact quarantine process as that from an individual initiating symptoms to their confirmation as a case, plus the delay for notified contacts to enter quarantine. The delay between symptom development and case confirmation will decrease with faster testing protocols, and indeed could become instant if presumptive diagnosis of COVID-19 based on symptoms were accepted in high-prevalence areas. The delay between contacts being notified and entering quarantine should be minimal with high levels of public understanding, as should the delay for case isolation. The efficacy of contact tracing (the *y* axis of Fig.3) can be identified with the square of the proportion of the population using the App, multiplied by the probability of the App detecting infectious contacts, multiplied by the fractional reduction in infectiousness resulting from being notified as a contact.

### Ethical considerations

Successful and appropriate use of the App relies on it commanding well-founded public trust and confidence. This applies to the use of the App itself and of the data gathered. There are strong, well-established ethical arguments recognizing the importance of achieving health benefits and avoiding harm. These arguments are particularly strong in the context of an epidemic with the potential for loss of life on the scale possible with COVID-19. Requirements for the intervention to be ethical and capable of commanding the trust of the public are likely to comprise the following. i. Oversight by an inclusive and transparent advisory board, which includes members of the public. ii. The agreement and publication of ethical principles by which the intervention will be guided. iii. Guarantees of equity of access and treatment. iv. The use of a transparent and auditable algorithm. v. Integrating evaluation and research in the intervention to inform the effective management of future major outbreaks. vi. Careful oversight of and effective protections around the uses of data. vii. The sharing of knowledge with other countries, especially low- and middle-income countries. viii. Ensuring that the intervention involves the minimum imposition possible and that decisions in policy and practice are guided by three moral values: equal moral respect, fairness, and the importance of reducing suffering (*28*). It is noteworthy that the algorithmic approach we propose avoids the need for coercive surveillance, since the system can have very large impacts and achieve sustained epidemic suppression, even with partial uptake. People should be democratically entitled to decide whether to adopt this platform. The intention is not to impose the technology as a permanent change to society, but we believe it is under these pandemic circumstances it is necessary and justified to protect public health.

## Discussion

In this study, we estimated key parameters of the SARS-CoV-2 epidemic, using an analytically solvable model of the exponential phase of spread and of the impact of interventions. Our estimate of R_0_ is lower than many previous published estimates, for example (*12*), (*29*), (*30*). These studies assumed SARS-like generation times; however, the emerging evidence for shorter generation times for COVID-19 implies a smaller R_0_. This means a smaller fraction of transmissions need to be blocked for sustained epidemic suppression (R < 1). However, it does not mean sustained epidemic suppression will be easier to achieve because each individual’s transmissions occur in a shorter window of time after their infection, and a greater fraction of them occurs before the warning sign of symptoms. Specifically, our approaches suggest that between a third and a half of transmissions occur from pre-symptomatic individuals. This is in line with estimates of 48% of transmission being pre-symptomatic in Singapore and 62% in Tianjin, China (*31*), and 44% in transmission pairs from various countries (*32*). Our infectiousness model suggests that the total contribution to R_0_ from pre-symptomatics is 0.9 (0.2 - 1.1), almost enough to sustain an epidemic on its own. For SARS, the corresponding estimate was almost zero (*9*), immediately telling us that different containment strategies will be needed for COVID-19.

Transmission occurring rapidly and before symptoms, as we have found, implies that the epidemic is highly unlikely to be contained by solely isolating symptomatic individuals. Published models (*9-11*) (*33*) suggest that in practice manual contact tracing can only improve on this to a limited extent: it is too slow, and cannot be scaled up once the epidemic grows beyond the early phase, due to limited personnel. Using mobile phones to measure infectious disease contact networks has been proposed previously (*34*-*36*). Considering our quantification of SARS-CoV-2 transmission, we suggest that this approach, with a mobile phone App implementing instantaneous contact tracing, could reduce transmission enough to achieve R < 1 and sustained epidemic suppression, stopping the virus from spreading further. We have developed a web interface to explore the uncertainty in our modelling assumptions (*25*). This will also serve as an ongoing resource as new data becomes available and as the epidemic evolves.

We included environmentally mediated transmission and transmission from asymptomatic individuals in our general mathematical framework. However, the relative importance of these transmission routes remain speculative based on current data. Cleaning and decontamination are being deployed to varying levels in different settings, and improved estimates of their relative importance would help inform this as a priority. Asymptomatic infection has been widely reported for COVID-19, e.g., (*14*), unlike for SARS where this was very rare (*37*). We argue that the reports from Singapore imply that even if asymptomatic infections are common, onward transmission from this state is probably uncommon, since forensic reconstruction of the transmission networks has closed down most missing links. There is an important caveat to this: the Singapore outbreak to date is small and has not implicated children. There has been widespread speculation that children could be frequent asymptomatic carriers and potential sources of SARS-CoV-2 (*38,39*).

We calibrated our estimate of the overall amount of transmission based on the epidemic growth rate observed in China not long after the epidemic started. Growth in Western European countries so far appears to be faster, implying either shorter intervals between individuals becoming infected and transmitting onwards, or a higher R_0_. We illustrate the latter effect in Figs *S*18 and *S*19. If this is an accurate picture of viral spread in Europe and not an artefact of early growth, epidemic control with only case isolation and quarantining of traced contacts appears implausible in this case, requiring near-universal App usage and near-perfect compliance. The App should be one tool among many general preventative population measures such as physical distancing, enhanced hand and respiratory hygiene, and regular decontamination.

An App-based intervention could be more powerful than our analysis here suggests, however. The renewal equation mathematical framework we use, while well adapted to account for realistic infectiousness dynamics, is not well adapted to account for benefits of recursion over the transmission network. Once they have been confirmed as cases, individuals identified by tracing can trigger further tracing, as can their contacts and so on. This effect was not modelled in our analysis here. If testing capacity is limited, individuals who are identified by tracing may be presumed confirmed upon onset of symptoms, since the prior probability of them being positive is higher than for the index case, accelerating the algorithm further without compromising specificity. With fast enough testing, even index cases diagnosed late in infection could be traced recursively, to identify recently infected individuals before they develop symptoms, and before they transmit. Improved sensitivity of testing in early infection could also speed up the algorithm and achieve rapid epidemic control.

The economic and social impact caused by widespread lockdowns is severe. Individuals on low incomes may have limited capacity to remain at home, and support for people in quarantine requires resources. Businesses will lose confidence, causing negative feedback cycles in the economy.

Psychological impacts may be lasting. Digital contact tracing could play a critical role in avoiding or leaving lockdown. We have quantified its expected success and laid out a series of requirements for its ethical implementation. The App we propose offers benefits for both society and individuals, reducing the number of cases and also enabling people to continue their lives in an informed, safe, and socially responsible way. It offers the potential to achieve important public benefits while maximising autonomy. Specific issues exist for groups within the population that may not be amenable to such an approach, and these could be rapidly refined in policy. Essential workers, such as health care workers, may need separate arrangements.

Further modelling is needed to compare the number of people disrupted under different scenarios consistent with sustained epidemic suppression. But a sustained pandemic is not inevitable, nor is sustained national lockdown. We recommend urgent exploration of means for intelligent physical distancing via digital contact tracing.

## Data Availability

All data are available in the manuscript or the supplementary material.

## Acknowledgments

We thank Will Probert, Andrei Akhmetzhanov, Alice Ledda, Ben Cowling, Gabriel Leung and Yuan Yang for helpful comments.

## Author contributions

Conceptualization: CF, DB. Data curation: LF, CW, AN, LZ. Funding acquisition: CF, MP. Investigation: LF, CW, MK, CF. Methodology: LF, CW. Visualization: LF, CW, MK, DB. Project administration: LA. Software: MK. Ethical analysis: MP, CF, DB. Writing, original draft: LF, CW, MP, DB, CF. Writing, review & editing: all authors.

## Data and materials availability

All data are available in the manuscript or the supplementary material. The code used for our analyses is publicly available at (*45*).

## Supplementary Materials

Materials and Methods

Figures *S*1-*S*

Tables *S*1-*S*

Supplementary text

Exponential growth rate

Inference of the distribution of generation times

Posterior probability of pre-symptomatic transmission

*β*(*τ*) and the renewal equation

Derivation of the impact of interventions

References (*40-44*)

